# Lead-OR: A Multimodal Platform for Deep Brain Stimulation Surgery

**DOI:** 10.1101/2021.08.09.21261792

**Authors:** Simón Oxenford, Jan Roediger, Luka Milosevic, Christopher Güttler, Philipp Spindler, Peter Vajkoczy, Wolf-Julian Neumann, Andrea Kühn, Andreas Horn

## Abstract

Deep Brain Stimulation (DBS) electrode implant trajectories are stereotactically defined using preoperative neuroimaging. To validate the correct trajectory, microelectrode recordings (MER) can be used to match the neuroanatomy with expected neurophysiological activity patterns, commonly using up to five trajectories in parallel. However, understanding their location in relationship to basal ganglia anatomy can be challenging. Here we present a tool that integrates resources from stereotactic planning, neuroimaging, MER and high-resolution atlas data to create a real-time visualization of the implant trajectory. We show a general correspondence between features derived from neuroimaging and electrophysiological recordings and present example use cases that demonstrate the functionality of the tool. The software toolbox is made openly available, extendable and holds translational potential in the field of stereotactic neurosurgery.

## Introduction

During Deep Brain Stimulation (DBS) surgery, different sources of information are used to ensure precise placement of the electrodes within the target structure. Functional stereotactic coordinates (defined relative to anatomical landmarks) are often used as a starting point (indirect targeting). Then, more importantly, preoperative MRI sequences optimized to visualize target structures are used to refine the initial plan (direct targeting). Surgical planning is usually carried out after fusing the MRI sequences with a computed tomography (CT) volume taken with the stereotactic frame already mounted to the cranium. The frame includes markers that are used to convert stereotactic coordinates (established in the planning software) to frame coordinates (applicable to mechanically adjust the stereotactic frame) in order to place electrodes to the intended target.

During the surgical procedure, microelectrode recordings (MER), as well as test-stimulations carried out using macroelectrodes are often used as an additional confirmation step of placement in the intended target site. While the necessity of the former step has been debated (***Aviles-Olmos et al., 2014***) and the procedure may lead to slightly increased rates of complications (***Zrinzo et al., 2012***), the experience of our own high-volume center is that roughly, every fifth patient’s surgical plan will be slightly altered based on electrophysiological signals, with similar experiences reported by others (***Lozano et al., 2019***). Of specific relevance is the role of brain shift occurring due to air entering the skull during surgery: even with optimal imaging and meticulous surgical planning beforehand, brain shift may lead to nonlinear displacement of the brain relative to the skull and stereotactic frame (***Halpern et al., 2008***) which can only be monitored intraoperatively (using the electrophysiological data recorded with microelectrode probes). While most centers analyze MERs by visual and auditory inspection from expert electrophysiologists, the first FDA and CE-approved machine-learning algorithms that facilitate this monitoring step have been recently introduced, for instance in form of the HaGuide system created by the company Alpha Omega (Alpha Omega Engineering, Nazareth, Israel; ***Thompson et al. (2018***)).

Still, even for experts, understanding and communicating the complex neuroanatomical and neurophysiological relationships within the clinical team during the procedure remains a major challenge. To account for this, Krüger et al. introduced the concept of Navigated Deep Brain Stimulation Surgery, showing that a visual feedback of the microelectrode position can be helpful to mentally envision the ongoing 3D scene (***Krüger et al., 2020***).

In parallel, reconstructions of DBS target regions based on elaborate MRI sequences have become increasingly precise (***Horn, 2019***; ***Krauss et al., 2021***). Specialized MRI sequences have been introduced to maximize visibility and boundary definitions of pallidal, thalamic (***Tourdias et al., 2014***; ***Sudhyadhom et al., 2009***; ***Vassal et al., 2012***) and subthalamic (***Santin et al., 2017***; ***Wang and Liu, 2015***) targets. But even when relying on a set of standard sequences (e.g., T1 and T2), modern reconstruction pipelines have the capability to reconstruct the STN and GPi with a precision that rivals manual expert segmentations (***Ewert et al., 2019***). Over recent years, these methods have made it possible to transform the 2D representations of stereotactic imaging slices into 3D models that are not only graphically appealing but indeed realistic and meaningful (***Horn and Kühn, 2015***). As a by-product, these tools have made it possible to accurately register atlas data into the patient-specific model. With *atlas data*, here, we refer to an array of ultra-high resolution imaging resources that could be based on histology (***Ilinsky et al., 2018***; ***Ewert et al., 2017***; ***Amunts et al., 2013***), postmortem MRI (***Edlow et al., 2019***) or even expert anatomical knowledge aggregated in three-dimensional fashion (***Petersen et al., 2019***). Similarly, atlas data could represent optimal stimulation sites defined on a group level, for instance in form of probabilistic sweet spot targets (***Dembek et al., 2019***; ***Boutet et al., 2021***; ***Elias et al., 2020***; ***Horn et al., 2017***) or tractography-defined DBS target atlases (***Li et al., 2020***; ***Treu et al., 2020***; ***Al-Fatly et al., 2019***).

Here, we present an integrative approach to combine information derived from neuroimaging and neurophysiology in a joint visualization platform. First, we build on recent validations of subcortical normalization routines to introduce a method to refine 3D models of subcortical targets on a single patient level. Second, we port our methodology for postoperative electrode localization established within Lead-DBS software (www.lead-dbs.org; ***Horn and Kühn*** (***2015***)) to the pre- and intraoperative realm, i.e. the one of stereotactic planning, microelectrode recordings and intraoperative testing. To achieve this, we present and validate a novel unified software framework termed Lead-OR that incorporates the following resources into a live visualization scene: (I) patient specific imaging, (II) stereotactic planning information, (III) live micro electrode positioning, (IV) MER feature extraction and (V) high resolution atlas imaging data. The capability of the system to integrate electrophysiological information with imaging data is explored in-depth. Beyond this feature, the tool also includes the possibility to visualize test-stimulations and live fiber tractography. The software framework is made available as an open source package (https://github.com/netstim/SlicerNetstim) and currently supports integration with the BrainLab Elements (Brainlab AG, Munich, Germany) planning software and a direct interface to the NeuroOmega system (Alpha Omega Engineering, Nazareth, Israel). Further integrations with other systems are planned, in the future.

## Methods

### Ethics statement

Lead-OR is intended for purely academic research use and does not have any form of government body regulatory approval. As such, any use of Lead-OR is strictly limited to Institutional Review Board (IRB) approved research studies at individual academic institutions. The collection and analysis of all patient data used for this article was approved by the Local Ethics committee of Charité — Universitätsmedizin Berlin (master vote EA2/145/21).

### Implementation environment

The tools used in this study are implemented in form of a 3D Slicer (Slicer) (***Fedorov et al., 2012***) extension (https://github.com/netstim/SlicerNetstim). The main module of the SlicerNetstim extension is Lead-OR, which assembles the different sources of information, as outlined in the following sections.

### Coordinate systems

The first step in aggregating data from different sources is to co-register their spatial relationship and coordinate systems (***Figure 1***). Lead-OR is based on Slicer’s world coordinate system (RAS). We use a linear transform to match the Head-Ring center and positive axes to the origin of this world-coordinate system. The planned central trajectory is then defined based on target coordinates, mounting type and Ring and Arc angles. The other trajectories are defined relative to the central one, following the configuration of the Ben-Gun microarray. As mentioned, currently, support for the NeuroOmega setup has been implemented, which uses a Ben-Gun array first introduced by Alim-Louis Benabid (***Benazzouz et al., 2002***).

**Figure 1.**
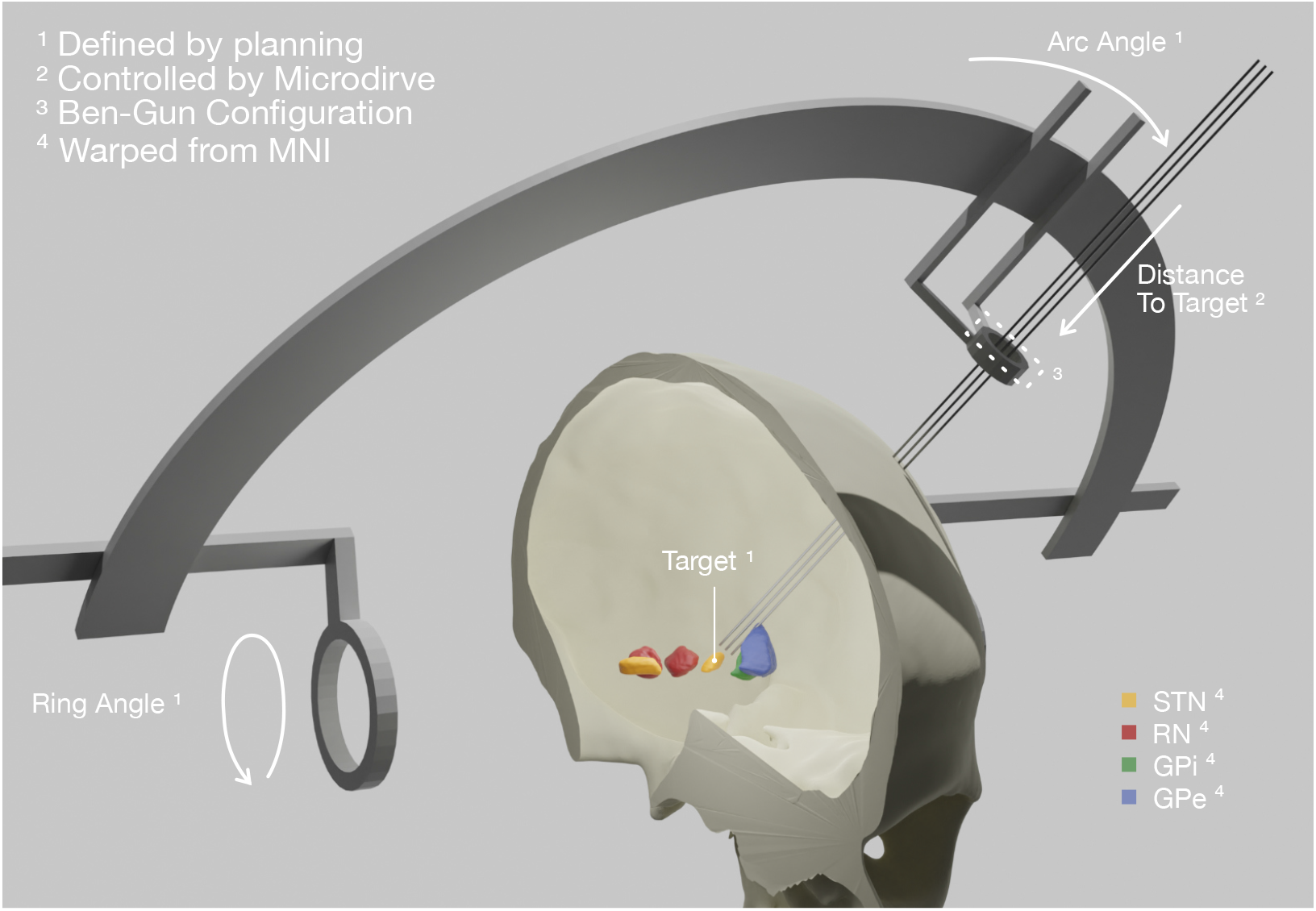
Patient-specific visualization generated by aggregating different sources of data. The stereotactic planning procedure defines the surgical target coordinate, as well as ring and arc angles, which together describe the central trajectory. The Ben-Gun configuration presented in the figure shows additional posteromedial and anterolateral trajectories, 2 *mm* apart from the central one. Up to five trajectories are currently supported by the software. In our current setup, the distance to the target is controlled by the NeuroOmega system, accessed with its SDK — but can alternatively be set manually within the tool itself. Relevant subcortical nuclei have been warped to patient space via a manually refined normalization.

These trajectories describe a line in space through which the macro, micro and definitive DBS electrodes are inserted. The last parameter to fully define their position varies throughout surgery, namely the distance to the planned target. This parameter is set by the Microdrive, allowing to move the electrodes along the trajectories while recording from the tip of the microelectrodes. In our current setup, this value is queried via the NeuroOmega Software Development Kit (SDK) and alternatively can be manually controlled within the software itself. Interfacing to similar systems as the NeuroOmega device will be possible given the open-source nature of our tool and creating such interfaces with other systems is in our interest, for the future.

To coregister the patient’s images and the frame reference, the tool uses a set of fiducial points defined in both coordinate systems (image and frame) which we extract from the surgical planning coordinates. Specifically, the anterior and posterior commissures (AC & PC) as well as a midsagittal point (MS) are used to create the transform (implemented using the fiducial registration module available within Slicer). Currently, an interface with the Brainlab Elements (Brainlab AG, Munich, Germany) stereotactic planning software is implemented (via PDF export in Elements and automated import in Lead-OR). Again, support for alternative planning tools is planned, for the future.

Finally, we incorporate high resolution atlas resources into the patient specific visualization scene. For the present examples within the manuscript, we used nuclei from the DISTAL (***Ewert et al., 2017***) and MNI PD 25 histology atlases (***Xiao et al., 2017***) that were defined in MNI space (ICBM 2009b Nonlinear Asymmetric, ***Fonov et al. (2009***)). Similarly, we imported histological sections from the BigBrain atlas (***Amunts et al., 2013***) and fibertract definitions provided by the holographic basal ganglia pathway atlas (***Petersen et al., 2019***). In the same fashion, virtually any type of atlas data could be imported to the patient scene, but it is crucial that this registration is of utmost precision. To account for this, we built on the long-standing methods development within Lead-DBS (***Horn and Kühn, 2015***; ***Horn et al., 2019***; ***Ewert et al., 2019***; ***Vogel et al., 2020***; ***Edlow et al., 2019***) but drastically extended the procedure with a novel manual refinement method, termed WarpDrive. Namely, an initial deformation field was calculated via a multispectral four-stage normalization step using the symmetric normalization (SyN) transformation model implemented within Advanced Normalization Tools (ANTs; http://stnava.github.io/ANTs/; ***Avants et al. (2008***)). This is implemented within the “effective: low variance + subcortical refinement” preset defined in Lead-DBS, which has been optimized for normalization of subcortical structures (***Horn et al., 2019***) and has shown to yield accurate segmentations of subcortical nuclei that rival the ones carried out by manually experts (***Ewert et al., 2019***; ***Vogel et al., 2020***). The deformation fields derived from this automated step are then further manually refined using a novel tool that is described in the next section.

### Normalization refinement

While normalization algorithms have become increasingly accurate (***Vogel et al., 2020***; ***Ewert et al., 2019***), their precision isn’t always perfect in single subjects and shows varying accuracy throughout the brain. Indeed, accurate automated registration of the basal ganglia nuclei presents a challenge to intensity-based registration methods given their low contrast between regions (***Ewert et al., 2019***).

Using our novel tool, WarpDrive, an experienced user can recognize such mismatches included in the automated normalization and manually refine the displacement field using point-to-point, line-to-line fiducials as well as a smudge tool. Manually entered fiducials are fed into the Plasti-match software (***Sharp et al., 2010***) (accessed as a command line module from within Slicer). Details about the WarpDrive tool will be reported and evaluated elsewhere. However, ***Figure 2*** shows an example of a manually refined normalization and ***Figure 2***–***video 1*** shows a demo application of the tool to refine atlas-to-patient fits in a surgical case.

**Figure 2.**
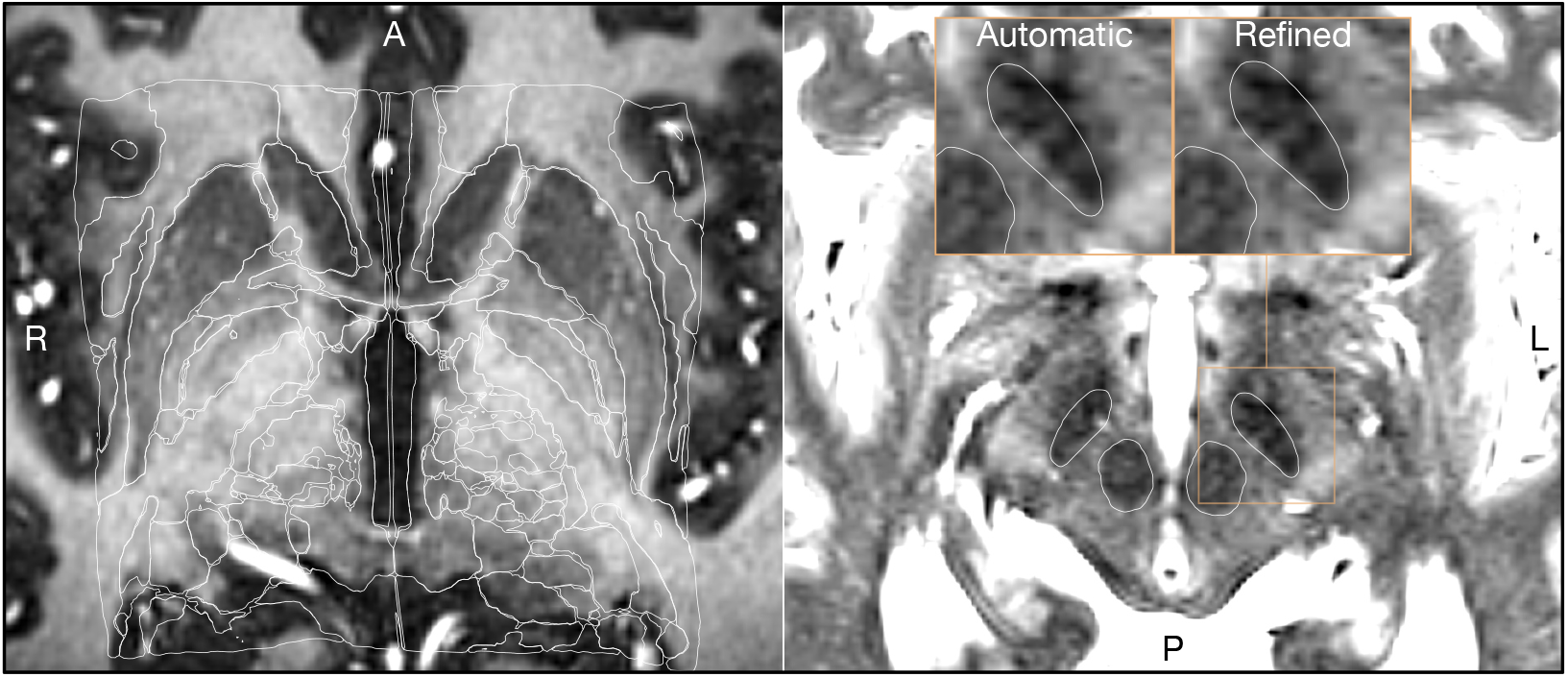
Output of a Lead-DBS / ANTs based automated normalization with and without subsequent manual refinement. Two MRI modalities are shown AC-PC aligned: FGATIR (left) and T2 (right). Both MRI modalities (together with T1, not shown here) were used as an input to the normalization step implemented in Lead-DBS, which allows multispectral registration using ANTs. The white outline shows atlases: MNI PD 25 histology (***Xiao et al., 2017***) (left) and DISTAL (***Ewert et al., 2017***) (right), both included within Lead-DBS. **Figure 2–video 1**. General overview of the visualizations and tools made available through the WarpDrive module implementation in Slicer. The video shows an image normalized to MNI space and how the Smudge and Drawing tools can help refine the displacement field.

### Real-time implementation

Lead-OR has the potential to be used in real-time during surgery. As mentioned above, one aspect of this is the continuous/live updating of the microelectrode distance to the surgical target while keeping the scene (i.e. multiple 2D and 3D views) synchronized. The interface to the NeuroOmega provides live data about how distant the microelectrodes are to the target, and also streams out electrophysiological recordings made in a real-time manner. Finally, test stimulations can be visualized including a function for live-tractography visualization estimating “activated” or “modulated” tracts.

To make this possible we included the NeuroOmega C++ SDK as part of a Slicer loadable module. This module sets up the connection to the NeuroOmega device and queries the distance to the target in specified time intervals. It also displays the available channels from which recorded electrophysiological data can be streamed, stored, processed and visualized.

Through the Lead-OR module the microelectrode Ben-Gun configuration is defined and the NeuroOmega channels are linked to the selected trajectories. Together with the aforementioned image-to-frame transform, as well as the distance to the target this allows to define the anatomical location of the electrophysiological signal in real-time. The features extracted from recordings are projected into the patient specific space and represented in the 2D/3D visualization (***Figure 3***).

**Figure 3.**
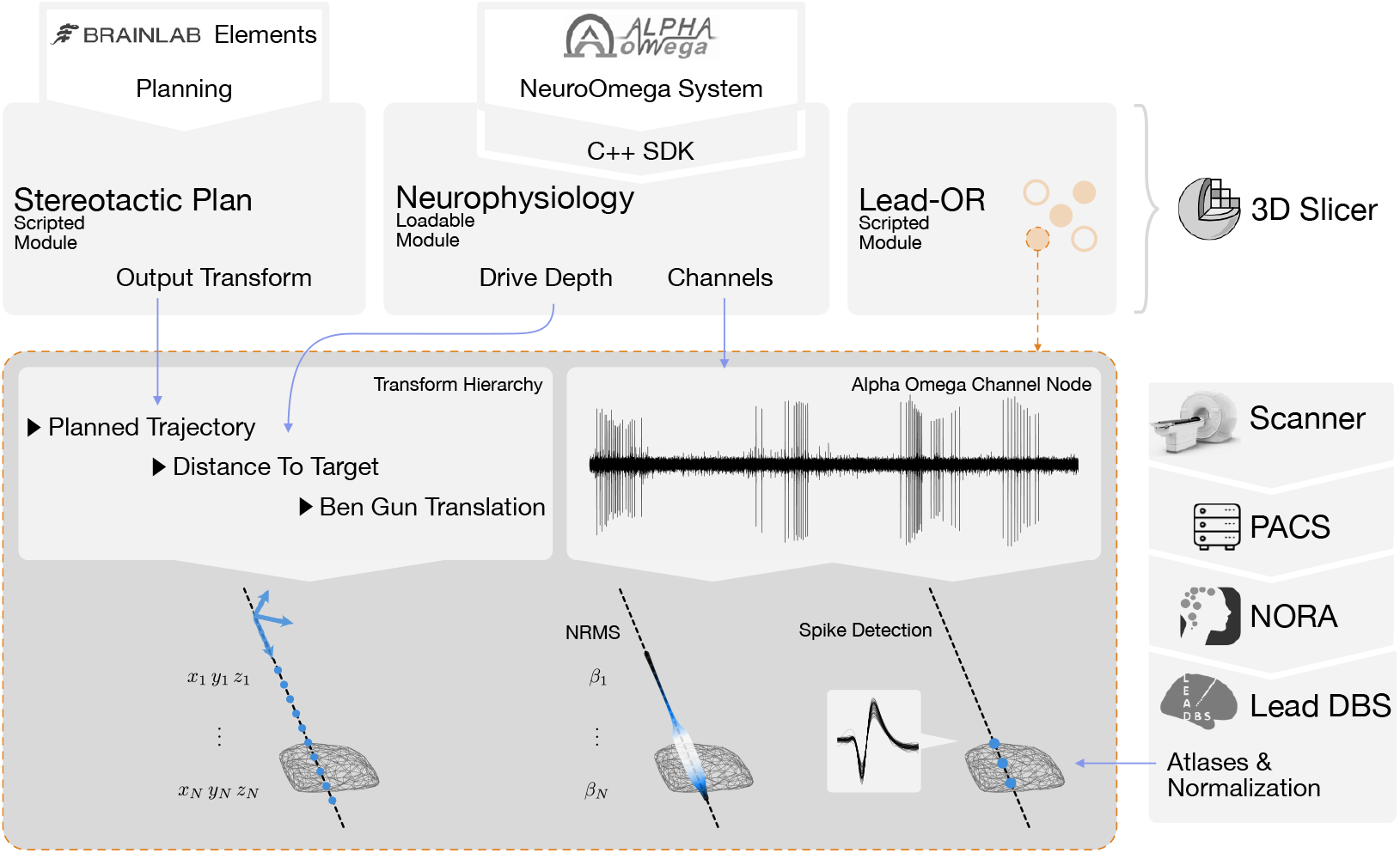
Overview of the SlicerNetstim Extension. The current setup shows interfaces with specific commercial products. Similar interfaces to competing tools are planned and will be included, in the future. A PDF plan exported from Brainlab Elements is used as an input to the Stereotactic Plan module to store the planned trajectory as a Slicer Transform. The NeuroOmega system is connected via its SDK through the Neurophysiology module, providing continuous updates about the drive depth and electrophysiological channel input. Finally, in the Lead-OR module, the Ben-Gun configuration is defined by selecting the used trajectories and assigning them to input channels from the NeuroOmega device. Using a transform hierarchy, the spatial position of the microelectrode is defined: the Ben-Gun translation is transformed by the distance to the target, this one itself being transformed by the planned trajectory. By doing so, the features extracted from the respective MER recordings can be mapped to their spatial location. At our center, an automatic pipeline for preprocessing data retrieved from a PACS system is set up using the NORA medical imaging platform (https://www.nora-imaging.com/) to automatically run the core Lead-DBS pipeline once images arrive in the hospital’s PACS system. This part (right-hand side) is not discussed in detail since largely specific to our center.

(Re-)developing a signal processing pipeline for electrophysiological data was not the focus of the present study since a multitude of tools exist, which could be integrated into Lead-OR, in the future. However, to demonstrate live-processing and -visualization of electrophysiological features, for now we included two minimal processing pipelines for MER. (Currently, no pipeline for local field potential recordings is included but this could be similarly extended given the open-source nature of the tool).

The first is the signal’s Normalized Root Mean Square (NRMS) value, which is computed as described in (***Zaidel et al., 2009***). For each step (Microdrive position), a stable part of the recorded data is extracted to compute the RMS on (see Zaidel et al. supplementary material). To obtain a normalized measure, the values along the trajectory are divided by the median of the first five stable steps. To visualize them in space we use a tube along the trajectory with varying radius and color — both redundantly representing NRMS magnitude. Potentially, in the future, radius and color could be assigned to represent different features that could graphically combine information derived from MER and LFP signals.

The second processing pipeline is based on spike analysis. This is implemented by running the WaveClus (***Chaure et al., 2018***) automatic pipeline with negative threshold on the recorded files once the drive moves to the next position. Clusters with less than one hundred spikes or in which 10 % of the Inter Spike Intervals (ISI) are below 3 *ms* or in which Signal to Noise Ratio (SNR) is less than 1.5, are discarded. SNR is computed as described in (***Joshua et al., 2007***) using the residual method. We assume the recordings capture Singe Unit Activity (SUA) instead of Multi Unit Activity (MUA), and thus each recording can represent none or one cluster of spikes. One of the reasons behind this assumption is, for example, that changes in amplitude recording from the same unit might be miss classified as different clusters. Spike clusters are represented as fiducials placed in the position they were detected. ***Figure 3*** summarizes the described live-processing setup.

### Stimulation module

Intraoperative assessment of stimulation-induced therapeutic as well as side effects can yield important information about electrode placement. For example, electrode placement close to the internal capsule may lead to tonic muscle contractions at low stimulation amplitudes. Often, these thresholds are intraoperatively identified by stimulating at increasing steps until muscle contraction and/or electromyography (EMG) activity is observed. Since Lead-OR already visualizes the patient specific location of the stimulation sites, volumes of tissue activated (VTA) could be used as seeds for tractography. Fiber analysis was carried out by accessing the logic of the SlicerDMRI module (***Norton et al., 2017***).

Obtaining preoperative diffusion MRI data is not part of clinical practice at all DBS centers. In cases where patient-specific dMRI data is not available, an alternative is to use normative fibers that are defined in template space and have been warped into patient space (similarly to atlas data). This process can at times even come with advantages, for example, the absence of false positive fibers when using manually curated normative datasets (***Petersen et al. (2019***); for a more thorough discussion see ***Horn and Fox*** (***2020***)). For the purpose of the present manuscript, we will refer to the term *tractography* the process to filter and visualize tracts derived from normative datasets or whole brain tractography connectomes (***Reisert et al., 2011***) that intersect with a region of interest (ROI) (in our case, the VTA).

To estimate the VTA, we use the simplified method proposed by Dembek, which defines the radius of a sphere based on stimulation amplitude and pulse width (***Dembek et al., 2017***). Varying values of from 0.5 to 1.0 were set for the constant *k*_2_ in Dembek’s formula (see supplementary material of the paper by Dembek et al. for the explanation of the parameter). In our present example, a value of ∼ 0.8 seemed to yield results that matched the recorded EMG data (***Figure 6***). Currently, this part of our manuscript should be seen as exploratory and stating a feasibility example only. Data to validate the approach on a larger number of patients beyond the present case was lacking. Further studies are needed to titrate the *k*_2_ value on a group level and to validate the stimulation module of Lead-OR, in general.

### Patient cohort and surgical procedure

Up to this point, we described the live-setup of Lead-OR. We aimed at evaluating accuracy of this setup by comparing imaging and electrophysiology derived markers on a group level. To do so, we retrospectively gathered data from patients that underwent DBS and processed it in a similar fashion as the real-time application. 52 patients were retrieved from cases undergoing STN-DBS surgery at Charité — Universitätsmedizin Berlin from 2017 until 2021.

Patients underwent bilateral DBS surgery targeted at the subthalamic nucleus. Surgery was either performed awake or under general anaesthesia. In case of the latter, the depth of narcosis was reduced before MERs to reduce potential effects of anaesthetic drugs.

The NeuroOmega System (Alpha Omega Engineering, Nazareth, Israel) was used with 2 to 5 microelectrodes in orthogonal (0^°^) or rotated (45^°^) Ben-Gun configuration to acquire MERs. Recordings were carried out from 10 *mm* above to 4 *mm* below the target with step sizes between 0.2 *mm* and 0.5 *mm* (with some exceptions common to clinical practice). Then, microelectrodes were removed and test-stimulations were applied at multiple heights above the target via macroelectrodes on central and alternate trajectories. Stimulations were done at increasing amplitude steps of 0.5 *mA* until identifying permanent side-effects. Additionally, therapeutic stimulation effects were evaluated when the surgery was performed in the awake state. Patients who underwent general anesthesia received additional EMG using needle electrodes to evaluate motor unit activity of eight muscles as indicator for activation of corticobulbar and corticospinal tracts. Finally, based on imaging, electrophysiological and clinical findings the surgical team decided upon the optimal depth and trajectory for permanent electrode implantation.

Of the 52 patients included in this study, 4 patients were discarded based on poor imaging quality and 16 based on poor electrophysiology signals (both determined by visual inspection). Additionally, taking the same considerations, 4 left and 4 right hemispheres were also discarded based on a low quality of electrophysiology data. MERs were saved as segments for each distance to the target value. Segments were discarded if they were contaminated by artifacts, or when their recording length was less than 4 seconds. With these considerations, we analyzed a final cohort of 32 patients (56 hemispheres) with a total of 236 trajectories.

### Imaging and electrophysiology processing

Pre- and postoperative imaging data were co-registered and normalized using Lead-DBS (***Horn et al., 2019***) followed by visual inspection and, if necessary, refinement using WarpDrive. The definitions of the central trajectories were extracted from stereotactic planning reports and the Ben-Gun configuration from recordings files. We computed the normalized root mean square of all trajectories and re-sampled them on a linear space with 0.1 mm distance to target resolution. Spike clusters were computed as described above. As mentioned, if more than one cluster was detected in a segment and satisfied the stated conditions, this was still considered a single unit activity (and represented as one cluster in further analysis).

Each recording segment had its own patient specific distance to target measure. In order to carry out group analyses, we defined a normalized distance to target. With the nonlinear deformation displacement fields, a link between the location of the trajectory and the ICBM 2009b NLIN ASYM (***Fonov et al., 2009***) (“MNI”) space was established. We then took a reference point in each trajectory computed as the nearest point to the STN target coordinates in MNI space from ***Caire et al. (2013***). The normalized distance to the target was defined by aligning the references of all trajectories. The alignment was done by displacing each trajectory by its reference position minus the average displacement from all trajectories. Furthermore, by using the link to MNI space we were able to compute the trajectory’s distance to the STN and the STN entry and exit sites (henceforth referred to as imaging defined STN boundaries). To do this, we used the STN as defined by the DISTAL atlas.

All spike clusters were mapped to the left hemisphere of the MNI space (right hemisphere co-ordinates were nonlinearly flipped). Then, we created an image of 0.22 *mm* isotropic resolution where each voxel represented the number of clusters detected divided by the number of segment recordings within 1 *mm* of the voxel’s center. This resulted in a cluster density volume in MNI space.

## Results

The main result of this work consists in an integrated software framework that links electrophysiological with imaging derived data within the same patient specific coordinate space during surgery. ***Figure 4*** shows the software output for a single case example including different forms of visualization and an exemplary match between DBS imaging and electrophysiology. ***Figure 4***–***video 1*** shows the live application of the tool in action.

**Figure 4.**
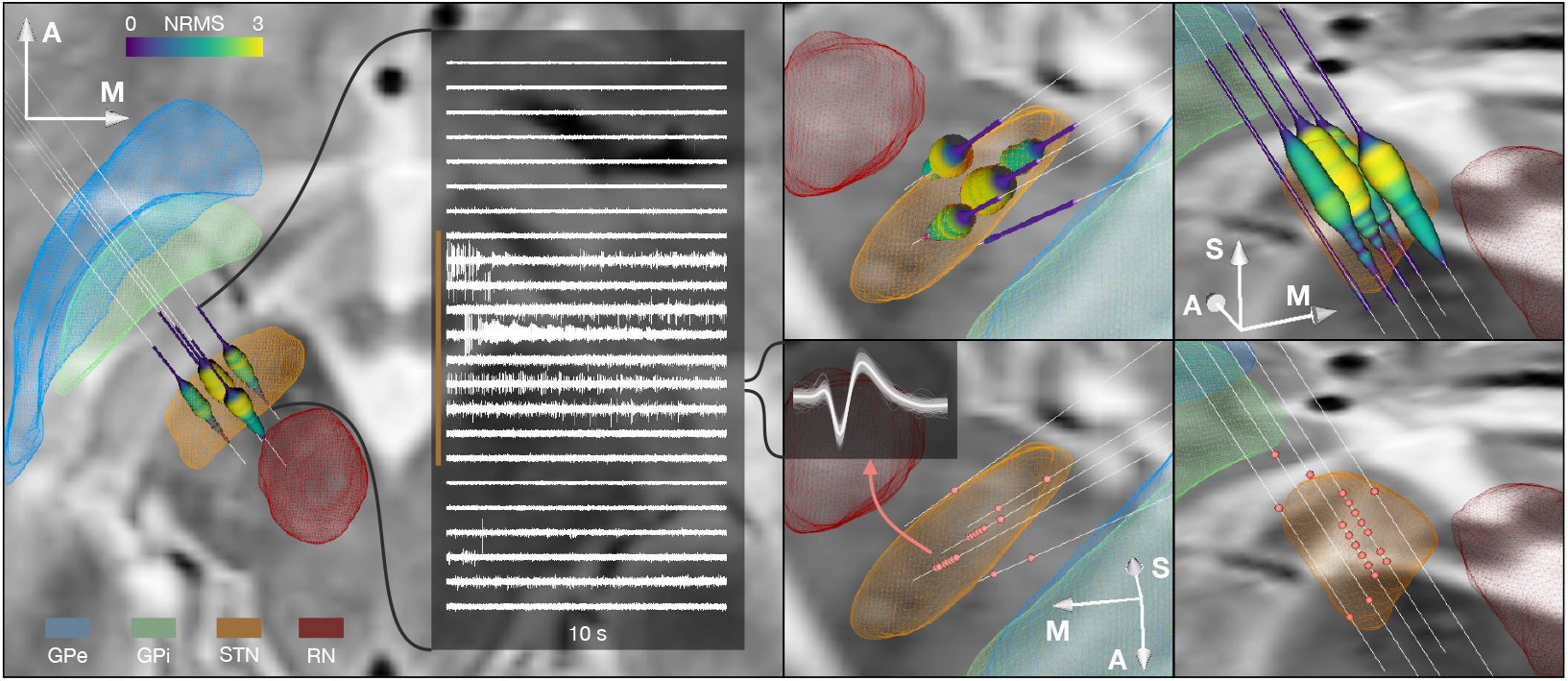
Example case showing trajectories, MER features and DISTAL atlas volumes mapped to patient space. 10 second recording snippets from one trajectory are displayed. Normalized root mean square (NRMS) activity is represented by a tube with varying diameter and color matching the value. Spike clusters, are represented by red point fiducials. **Figure 4–video 1**. General overview of the Lead-OR real-time application. The video shows patient specific imaging together with atlases, the BigBrain template (***Amunts et al., 2013***), planned microelectrode trajectories, MERs and feature extraction and a test-stimulation example.

***Figure 5*** shows the 236 trajectories retrospectively gathered from 32 patients, arranged from left to right based on their distance to the STN and vertically aligned with the normalized distance to target. Electrophysiology traces were plotted with STN entry and exit markers derived from imaging. Comparing the NRMS from the bottom 20 % (outside of the STN) to the top 20 % revealed an anatomical region with significant differences (*p* < 0.01) within the imaging defined STN boundaries (defined as the median of the top 20 % STN boundaries). In other words, NRMS values inside this part of the STN were significantly higher than outside of it. Data were compared using nonparametric Wilcoxon’s signed rank test and multiple comparisons were corrected using False Discovery Rate (FDR) (***Benjamini et al., 2006***).

**Figure 5.**
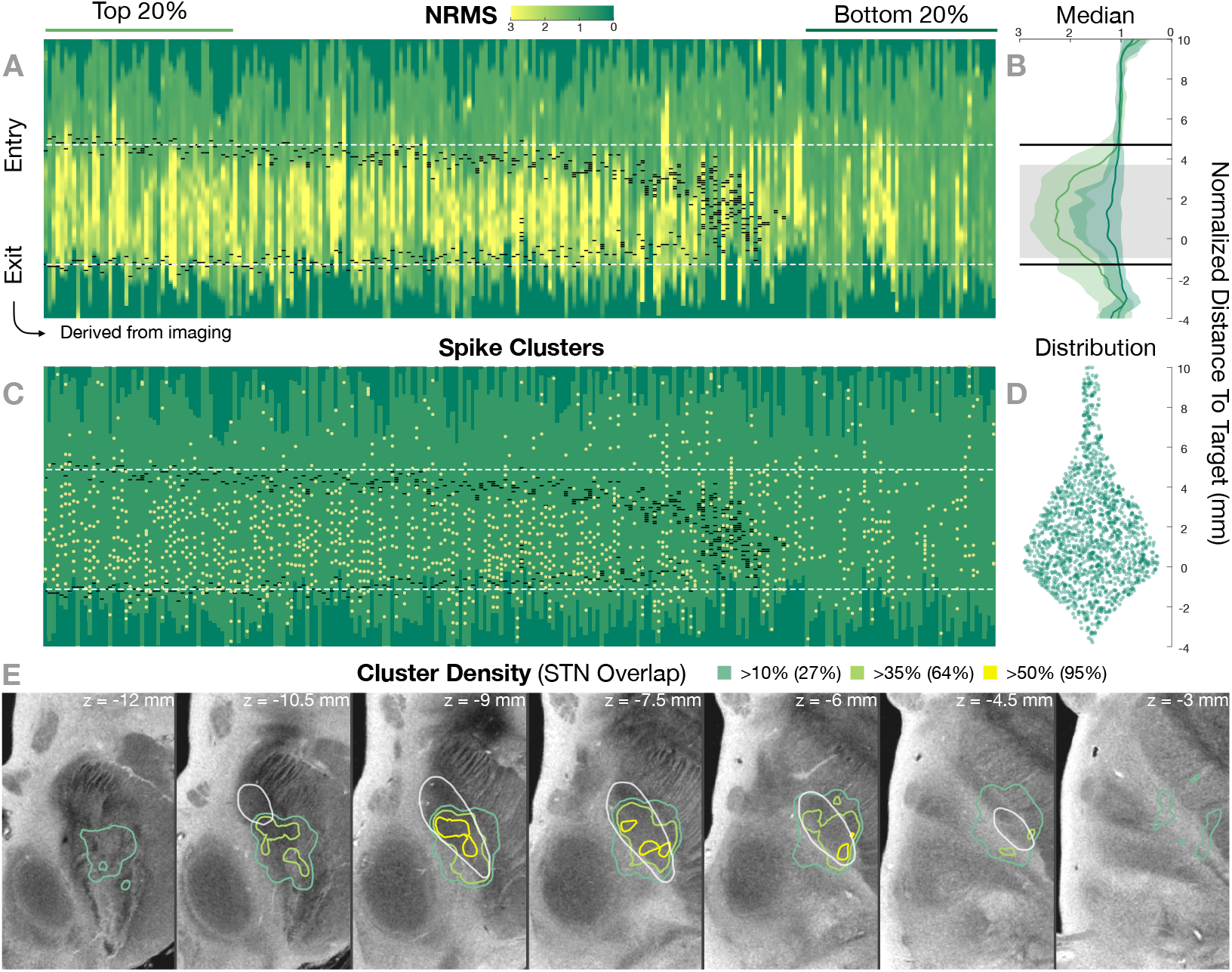
Retrospective group analysis investigating agreement between imaging- and electrophysiology-defined STN. In **A** and **C**, each trajectory is presented as a column, showing normalized root mean square activity (NRMS) and spike clusters, respectively, with the normalized distance to target denoted on vertical axes. Trajectories are sorted from left to right according to their distance to the STN as defined in the DISTAL atlas (***Ewert et al., 2017***). Dark green values (indicating NRMS of zero) represent no recordings at these sites. Black dashes represent STN entry and exit, and the dashed white line the median entry and exit for the top 20 %. **B** shows comparisons between bottom and top trajectories, with the gray area representing a significant band (nonparametric Wilcoxon’s signed rank test *p* < 0.01 with FDR correction), which resides within the STN. The plots show median, 0.25 and 0.75 quantiles. **D** shows the overall distribution of spike clusters. **E** shows isosurfaces of a volume where each voxel contains the number of clusters detected divided by the number of recordings carried out within 1 *mm* distance to the location (cluster density). The legend shows the percentage of the volume overlap with the STN at different thresholds. The 7 *T* MRI *ex vivo* human brain template (***Edlow et al., 2019***) is shown as the background image with DISTAL STN outline.

The cluster density volume in MNI space also showed a general agreement with the imging-derived STN: when thresholding the volume based on increasing density values, the overlap with the STN region was higher (95 % overlap at a 50 % cluster density threshold; ***Figure 5***).

***Figure 6*** shows an example case using the test-stimulation setup with live volume activation tractography and corresponding EMG activity invasively recorded during surgical routine using a needle-electrode inserted to the brachioradialis muscle. We also refer to ***Figure 4***–***video 1*** for a demonstration of the real-time application of this module.

**Figure 6.**
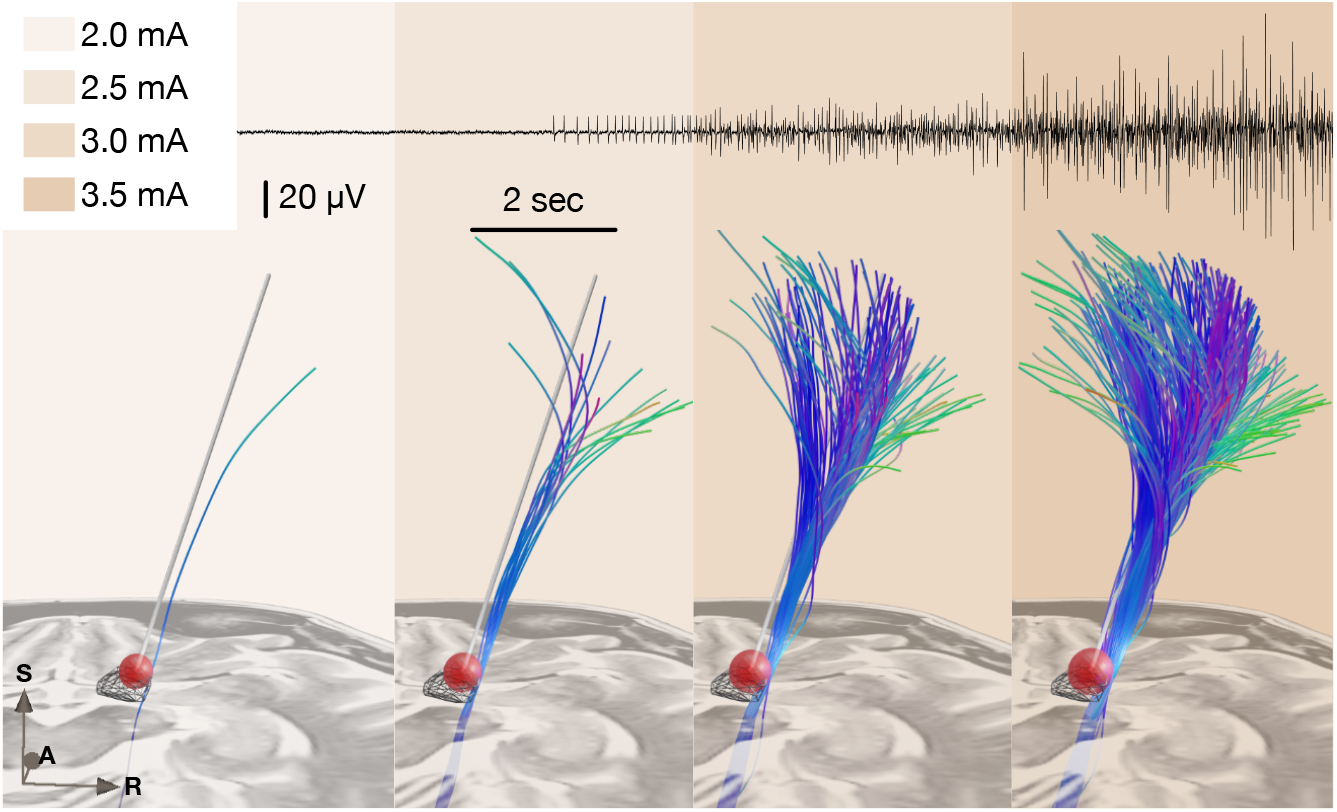
Example of test-stimulation setup (also see ***Figure 4***–***video 1*** for demonstration of a real-time application). A simplified stimulation volume is modeled based on the applied test stimulation parameters following the approach of ***Dembek et al. (2017***). From a set of pre-defined fibertracts representing the internal capsule (without hyperdirect components; ***Petersen et al. (2019***)) that were registered to patient space, fibers passing through the volume were visualized in real-time. Alternatively, tractograms obtained based on diffusion MRI of the individual patient data or normative connectomes could be used. The top panel shows needle EMG activity that was recorded within clinical routine from the brachioradialis muscle during stimulation in the same patient. Colors represent stimulation amplitude. After a preliminary exploratory analysis of the *k*_2_ parameter from Dembek’s formula, a value of 0.8 was used for the shown example.

## Discussion

Multiple take-home points can be drawn from this study. First, we established a software pipeline to integrate imaging and electrophysiology results within an interactive real-time application during DBS surgery. The setup interacts with commercial tools for surgical planning and MERs and has the capability to visualize and analyze data in various forms. While currently, a fixed set of interfaces to commercial tools is available, the open-source nature of the software will allow integration of links to other platforms. Second, atlas data from ultra-high resolution resources may be integrated into the tool. For instance, whole-brain histological atlases, such as the BigBrain dataset (***Amunts et al., 2013***) or stereotactic 3D atlases, such as the DISTAL (***Ewert et al., 2017***) or Human Thalamus Atlas (***Ilinsky et al., 2018***) could be integrated. In a way, these atlases would “replace” commonly used histological reference atlases available in book format, such as the Schaltenbrandt-Wahren (***Schaltenbrand et al., 1977***) or Talairach atlases (***Talairach and Tournoux, 1988***). While these book resources have been and still are invaluable to the field, they lack the possibility to be deformed into native patient space and to be digitally represented in direct synopsis with patient imaging and electrophysiology. Instead, whole-brain resources will grow in number, resolution, and quality in the foreseeable future (***Horn, 2019***; ***Krauss et al., 2021***; ***Sui et al., 2021***; ***Vedam-Mai et al., 2021***). Similarly to anatomical atlas resources, optimal target definitions (“sweet spots”) or even connectomic/tract-based target definitions could one day be integrated to guide DBS surgery — after proper and prospective validation of such datasets and applied methods (***Dembek et al., 2019***; ***Boutet et al., 2021***; ***Elias et al., 2020***).

The tools, methods and software described here are not approved under any regulations and are not intended to assist in making clinical decisions. Rather, we present them for use in purely research driven purposes under proper IRB approval in study contexts. As such, they may be powerful to further explore the interplay between electrophysiology and imaging, to validate biophysical models and to better characterize patient specific data. We see special value in integrating MER-derived measures to the anatomical realm and in the integration with imaging findings. Our aim was to produce a set of use-cases each of which could open larger windows of opportunities for upcoming studies. For instance, we included two MER processing pipelines in the present study, which have been studied previously in different publications (***Koirala et al., 2020***; ***Boëx et al., 2018***; ***Zaidel et al., 2009***). The reason for their adoption was mostly demonstrative and we do not claim for them to be the best choices when it comes to studying STN activity. Future work involves analyzing differences in these and similar processing pipelines to derive a better understanding of MER physiology. Given the open-source nature of this project it will be feasible to extend usability and incorporate complementary approaches.

In a similar vein, we see larger potential in the field of activation tractography by studying stimulation spread across brain tissue (with biophysical models that could range around varying degrees of complexity (***Butenko et al., 2020***; ***Gunalan et al., 2017***; ***Howell et al., 2019***; ***Noecker et al., 2021***). Differences in connectomes (***Horn and Blankenburg, 2016***) vs. pathway atlases (***Petersen et al., 2019***; ***Alho et al., 2019***; ***Middlebrooks et al., 2020***) vs. individual tractography (***Akram et al., 2017***) acquired in the specific patient could be investigated directly within the operation theatre. We foresee that such studies could lead to a better understanding of the mechanism of action of DBS. The present study for now showcases this application of visualizing test stimulations in limited and anecdotical form (also see ***Figure 4***–***video 1***), warranting further investigation and validation.

Finally, we see large potential in the use and further aggregation of ultra-high resolution atlas data. Already, such datasets have been emerging and incorporated into DBS applications (***Edlow et al., 2019***; ***Horn et al., 2017***). However, we foresee additional datasets that may revolutionize our definition of anatomy and brain connectivity in the future. For instance, the Jülich group has announced an upcoming version of the BigBrain dataset (***Figure 7*** & ***Figure 4***–***video 1***) that will be available in 1 *μm* resolution (***Horn, 2021***). A recent normative diffusion-MRI connectome available in 760 *μm* resolution was based on a nine-hour long scan of a living human brain (***Wang et al., 2021***). Similarly, a structural brain template of the human brain available in 100 *μm* resolution was acquired by scanning a postmortem brain over 100 hours at 7 *T* (***Figure 7***) (***Edlow et al., 2019***). A recently published pathway atlas of the basal ganglia used expert knowledge and insights from animal studies to create the most realistic set of subcortical fibers available to date (***Petersen et al., 2019***). Similar applications involve histological mesh tractography — a novel technique to create accurate tract representations based on histological data (***Alho et al., 2021***) or expert-curated sets of fiber-bundles created by tractography on diffusion MRI data from 1,000 subjects (***Middlebrooks et al., 2020***). We foresee great use of such resources if the process of registering them to patient space is truly accurate. The WarpDrive method present here could embody a missing link in the evolution of making co-registration methodology as precise as possible — with specific focus to regions of particular interest (such as the DBS target zone in our application). For instance, if our aim was to overlay the BigBrain atlas to support our anatomical knowledge within and around the STN, it is of crucial importance that the registration between atlas and patient imaging of the STN area is meticulously precise. Instead, registration accuracy of, e.g., the parietal lobe, will be of lesser importance in this particular scenario. WarpDrive gives the user the necessary toolkit to realize highly precise warps, while focusing on specific regions of interest (***Figure 7*** and ***Figure 2***– ***video 1***).

**Figure 7.**
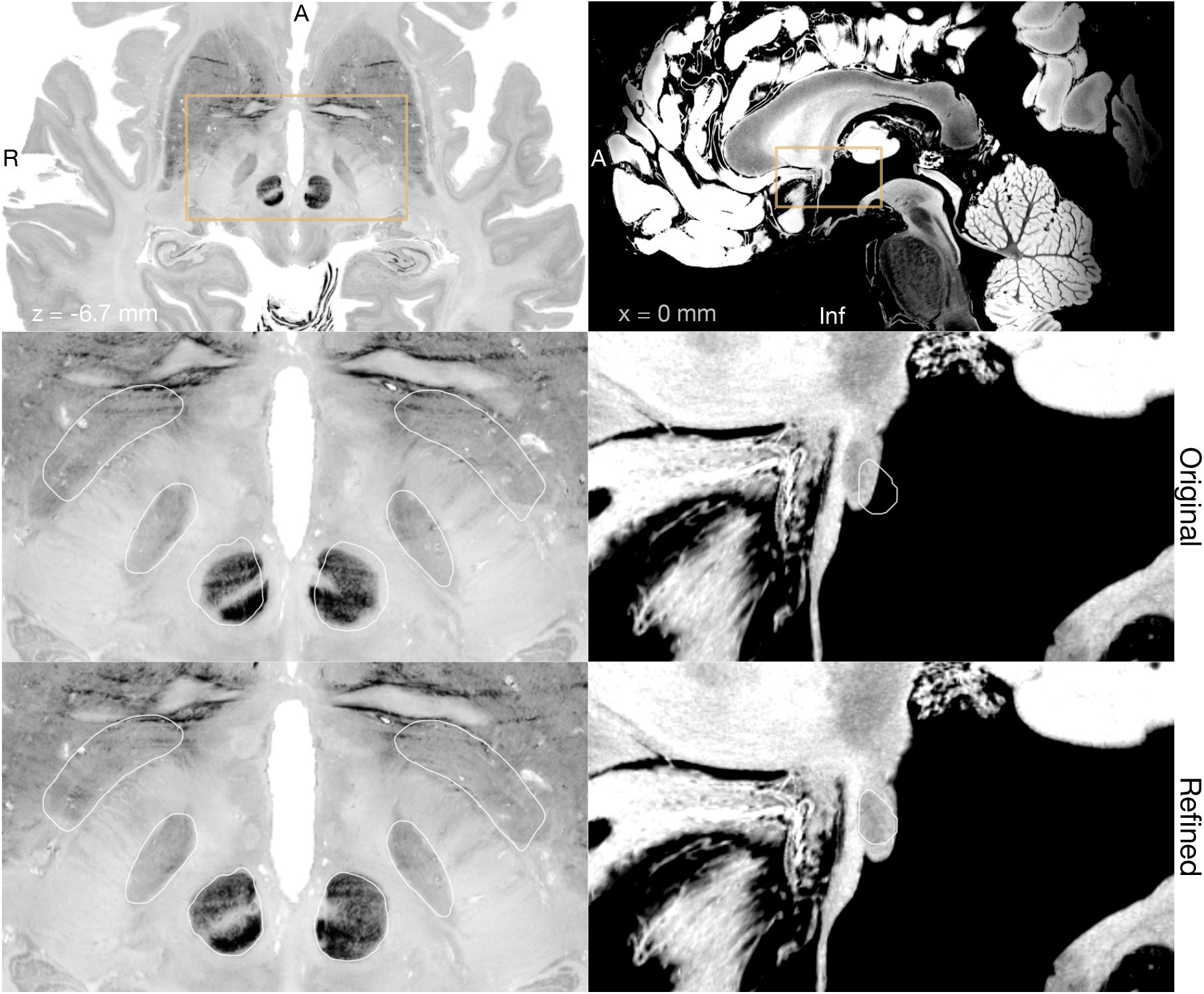
Big Brain (***Amunts et al., 2013***; ***Xiao et al., 2019***) (left) and 7 *T* MRI *ex vivo* human brain template (***Edlow et al., 2019***) (right) are two high resolution (100 *μm* isotropic) imaging resources that can be used from Lead-DBS and Lead-OR. The middle panel shows a close-up in a plane using the original transformation to MNI space (Xiao et al. in the case of Big Brain) with white outlines of DISTAL atlas (***Ewert et al., 2017***) (left) and Anterior Comissure from ***Neudorfer et al. (2020***) (right). The bottom panel shows the same slice, but using a refined transformation using Lead-DBS and WarpDrive.

Our results demonstrate general agreement between imaging and electrophysiology data on a group level. However, as can be seen in ***Figure 5***, the agreement is not 100 %. Namely, we can observe presence of activity and high neuronal density in some locations outside of our image-derived model of the STN and vice-versa (we observe no activity within voxels that form part of the STN). This emphasizes the possibility of a lack of congruence between preoperative imaging and intraoperative electrophysiological delineation of the STN. Some of these discrepancies could be explained physiologically, for example seeing activity from regions other than the STN (i.e., thalamic recordings that may be encountered dorsal to the STN, or recordings from substantia nigra ventrally). However, true mismatch of the two sources of information (imaging and electrophysiology) *in some cases* is indeed something we would *expect*. Namely, we should not forget that the tool is entirely designed to facilitate integration and visualization of *different* sources of information in parallel. If both would perfectly agree in each single case, there would be no need to acquire MER data in the first place. For instance, brain shift could have occurred, which is the main reason to pursue MER recordings in the first place. Hence, in cases where imaging and electrophysiology derived information do not match precisely, it will be up to the expert medical professional to integrate results and form their clinical decisions during surgery.

### Limitations

Other explanations for disagreement between imaging and electrophysiological data will directly inform limitations that apply to the current study. The occurrence of brain shift could be seen as a limitation but also as a feature of our approach (see above). However, true limitations may arise from imprecisions of the imaging pipeline itself. Although a dedicated multispectral imaging pipeline was applied (in form of Lead-DBS software), which has shown to create meaningful models of DBS in various studies, there will always be a certain degree of imprecision that is unavoidable when using imaging to segment subcortical nuclei. Here, we aimed to further minimize this imprecision by introducing the WarpDrive tool. However, a downside of this tool could be seen in the fact that it involves manual and observer-dependent steps. Detailed anatomical knowledge and optimal imaging quality are needed to achieve maximal registration accuracy. Ideally, multispectral sets of preoperative images that include specialized sequences optimized for the basal ganglia should be used (***Krauss et al., 2021***). Use of ultra-highfield (i.e., 7 *T*) imaging could represent a useful alternative (***Forstmann et al., 2017***), but in this case danger could arise from increased distortion artifacts exactly and especially in the center of the brain (***Neumann et al., 2015***). Hence, as in the procedure of DBS surgery itself, optimal imaging data quality and meticulous use of tools, as well as optimal levels of methodological insights are needed to assure safe and successful applications. Finally, the MER analysis also comes with limitations.

First, as the data was collected in retrospective fashion, durations of recordings and distances in recording steps when advancing towards the target were not exactly consistent throughout the whole dataset. Second, cardioballistic artifacts as well as gradual displacement of brain tissue leading to attenuation of spike amplitudes are recognized problems when applying spike-sorting algorithms, in general. Moreover, anesthesia and wakefulness of patients may have an impact on the recordable neurophysiological activity patterns and should be considered when making assumptions about the relationship between neuroanatomy and neurophysiology. While here, patients were awake in general, this followed periods of anesthesia. In the future, additional automatic EEG and EMG activity analysis could further augment the validity of the approach. In general, however, the main aim of this work was to demonstrate usage of the tool, while dedicated analyses investigating specific neuroscientific questions should take such nuances into consideration.

## Conclusion

We present a method and open-source software tool to visualize results derived from microelectrode recordings in anatomical space together with information derived from patient specific MRI data, as well as high resolution atlas resources during DBS surgery. We demonstrate general agreement between imaging and electrophysiology derived measures, as well as examples of unavoidable discrepancy between the two modalities. The tool has potential to empower scientific studies investigating several topics outlined in our discussion, as well as high potential for clinical translation and represents a first step to help integrate information across sources within two- and three-dimensional visualization scenes. While the software is not certified and intended for scientific use under IRB approval only, subsequent steps will involve improving and extending the different components of the software to achieve a reliable multimodal patient specific navigator capable of assisting clinical decision making.

## Data Availability

The DBS datasets generated during and analyzed within the current study are not publicly available due to data privacy regulations of patient data but are available from the corresponding author upon reasonable request. All code used to analyze the dataset is available within Lead-DBS /-OR software (https://github.com/netstim/leaddbs; https://github.com/netstim/SlicerNetstim).

## Acknowledgments

We would like to thank Alaa Hanna from AlphaOmega for methodological support in interfacing with the NeuroOmega SDK. While the present software implementation works with two commercial products for surgical planning and the intraoperative procedure, the choices of these systems were arbitrary (defined by what was present at our center) and do not mean any form of endorsement whatsoever. No industry funding was received to carry out this study.

Part of this study was presented and worked on during the 35th NA-MIC Project Week (***Kapur et al., 2016***). We would like to thank the NA-MIC community, specially to Dr. Andrass Lasso and Dr. Steve Pieper for the help and discussions on 3D Slicer modules implementation.

